# Maladaptive laterality in cortical networks related to social communication in autism spectrum disorder

**DOI:** 10.1101/2022.06.21.22276730

**Authors:** Andrew S. Persichetti, Jiayu Shao, Stephen J. Gotts, Alex Martin

## Abstract

Neuroimaging studies of individuals with autism spectrum disorders (ASD) consistently find an aberrant pattern of “reduced” laterality in brain networks that support functions related to social communication and language. However, it is unclear how the underlying functional organization of these brain networks is altered in ASD individuals. We tested four models of “reduced” laterality in a social-communication network in seventy ASD individuals (14 females) and a control group of the same number of tightly matched typically developing (TD) individuals (19 females) using high quality resting-state fMRI data and a method of measuring patterns of functional laterality across the brain. We found that a functionally defined social-communication network exhibited the typical pattern of left laterality in both groups, while there was a significant increase in laterality of homotopic regions in the right hemisphere in ASD individuals. Furthermore, left laterality was positively correlated with a measure of verbal ability in both groups, while right laterality in ASD, but not TD, individuals was negatively correlated with the same verbal measure. Crucially, these differences in patterns of laterality were not found in two other functional networks and were specifically correlated to a measure of verbal, but not visuospatial, ability. These results suggest that previous reports of “reduced” laterality in social-communication regions in ASD is due to a shift towards functional decoupling of the two hemispheres, which may cause them to act as independent systems with the atypical right-lateralized network being maladaptive.

## Introduction

Impaired social communication is a core behavioral phenotype across the autism spectrum, ranging from a complete lack of ability on the low end to subtle deficits in high-functioning individuals (Goldstein et al., 1994; Boucher, 2003, 2012; Rapin and Dunn, 2003; American Psychiatric Association, 2013). Consistent with these behavioral deficits, one of the most robust neuroimaging findings in (usually high functioning) individuals with autism spectrum disorders (ASD) is an aberrant pattern of “reduced” laterality of the brain networks that support functions related to social communication and language (Lindell and Hudry, 2013; Herringshaw et al., 2016). For example, functional MRI (fMRI) studies that use communication-and language-based tasks consistently find a reduced difference in the magnitude of neural responses between typically left-lateralized regions and homotopic regions in the right hemisphere in ASD compared to typically developing (TD) individuals (Herbert et al., 2002; Boddaert et al., 2003; Takeuchi et al., 2004; Wang et al., 2006; Harris et al., 2006; Kleinhans et al., 2008; Knaus et al., 2008, 2010; Redcay and Courchesne, 2008; Tesink et al., 2009; Anderson et al., 2010; Eyler et al., 2012; Jouravlev et al., 2020). While such results suggest that typically left-lateralized cortical networks that support social communication and language are more symmetrically distributed across hemispheres in ASD, hemispheric differences in task-based responses between ASD and TD groups could be due to several underlying changes to the functional organization of these brain networks. For example, atypical patterns of task responses in ASD could be due to an intact left-lateralized cortical network communicating more with the right hemisphere or a weakened left-lateralized network that results in compensatory activity in the right hemisphere. Therefore, it is necessary to measure patterns of functional connectivity within and across hemispheres to understand how brain networks underlying social communication are reorganized in ASD.

In TD individuals, left-lateralized networks that support functions requiring rapid cortical interactions, such as language, communication, and fine motor control, tend to communicate more exclusively within hemisphere than with regions in the right hemisphere (Semmes, 1968; Lackner and Teuber, 1973; Poeppel, 2003). By contrast, typically right-lateralized networks that support functions requiring integration of information across the hemispheres (e.g., the right-lateralized visuospatial attention network requires bilateral representations of space) also communicate strongly with left hemisphere regions (Corbetta and Shulman, 2011). To probe these distinct patterns of lateralization, a recent study developed two metrics of laterality based on patterns of resting-state functional connectivity: one that measures the degree of within-hemisphere communication from each location in cortex relative to the homotopic location in the other hemisphere, referred to as “Segregation,” and another that measures between-hemisphere communication, referred to as “Integration”. The authors found that the degree to which regions in a left-segregated frontotemporal network were left lateralized was positively correlated with a measure of verbal ability, while the degree to which regions in the right-lateralized visuospatial attention network were integrated with the left hemisphere was positively correlated with visuospatial ability (Gotts et al., 2013). In the current study, we use these validated laterality metrics to adjudicate between potential models of “reduced” left laterality in social communication regions in ASD.

## Methods

### Participants

Seventy individuals [mean (SD) age = 19 (3.8) years; 14 female] who met the DSM-V criteria for ASD (American Psychiatric Association, 2013), as assessed by a trained clinician, were recruited for this experiment. In addition, seventy individuals with no history of psychiatric or neurological disorders [mean (SD) age = 19.7 (3.7) years; 19 female] served as the TD control group. There was no significant difference between the ages of the two groups (t_(69)_=1.14, p=0.26). Subsets of the resting-state data from these individuals have been used in a number of our previous studies (e.g., Gotts et al., 2012a, 2013; Ramot et al., 2017a; Jasmin et al., 2019a; Power et al., 2019a; Persichetti et al., 2021). All participants gave informed consent under an NIH Institutional Review Board-approved protocol (10-M-0027, clinical trials number NCT01031407).

### Behavioral measures

The Wechsler Abbreviated Scale of Intelligence (WASI - Wechsler, 1999) was administered within 1 year of the scanning session to all participants in each group [mean (SD) Full-score IQ, TD: 116.1 (11); ASD 114.2 (12.9); t_(69)_=1.15, p=0.25]. We used the individual T-scores (normative mean = 50, SD = 10) from the vocabulary (Verbal IQ) and block design (non-verbal IQ) subtests in a correlation analysis with the neuroimaging data. These scores were missing from one participant in each group. We chose these subtests because they have been shown to have strong and selective associations with verbal/language and visuospatial abilities, respectively (Warrington et al., 1986; Gotts et al., 2013; Kenworthy et al., 2013).

### MRI data acquisition

Scanning was completed on a General Electric Signa HDxt 3.0 T scanner (GE Healthcare) at the National Institutes of Health Clinical Center NMR Research Facility. For each participant, T2*-weighted blood oxygen level-dependent (BOLD) images covering the whole brain were acquired using an 8-channel receive-only head coil and a gradient echo single-shot echo planar imaging sequence (repetition time, TR = 3500 ms, echo time, TE = 27 ms, flip angle = 90°, 42 axial contiguous interleaved slices per volume, 3.0-mm slice thickness, field of view, FOV = 22 cm, 128 × 128 acquisition matrix, single-voxel volume = 1.7 × 1.7 × 3.0 mm^3^). An acceleration factor of 2 (ASSET) was used to reduce gradient coil heating during the session. In addition to the functional images, a high-resolution T1-weighted anatomical image (magnetization-prepared rapid acquisition with gradient echo, or MPRAGE) was obtained (124 axial slices, 1.2-mm slice thickness, FOV = 24 cm, 224 × 224 acquisition matrix).

### fMRI procedure

During the resting scans, participants were instructed to relax and keep their eyes fixated on a central cross. Each resting scan lasted 8 min 10 s for a total of 140 consecutive whole-brain volumes. Independent measures of cardiac and respiratory cycles were recorded during scanning for later artifact removal.

### fMRI data preprocessing

All data were preprocessed using the AFNI software package (Cox, 1996). First, the initial 3 TRs from each EPI scan were removed to allow for T1 equilibration. Next, 3dDespike was used to bound outlying time points in each voxel within 4 standard deviations of the time series mean and 3dTshift was used to adjust for slice acquisition time within each volume (to t=0). 3dvolreg was then used to align each volume of the resting-state scan series to the first retained volume of the scan. White matter and large ventricle masks were created from the aligned MPRAGE scan using Freesurfer (e.g., Fischl et al., 2002). These masks were then resampled to EPI resolution, eroded by 1 voxel to prevent partial volume effects with gray matter voxels, and applied to the volume-registered data to generate white-matter and ventricle nuisance regressors prior to spatial blurring. Scans were then spatially blurred by a 6-mm Gaussian kernel (full width at half maximum) and divided by the mean of the voxelwise time series to yield units of percent signal change.

The data were denoised using the ANATICOR preprocessing approach (Jo et al., 2010). Nuisance regressors for each voxel included: 6 head-position parameter time series (3 translation, 3 rotation), 1 average eroded ventricle time series, 1 “localized” eroded white matter time series (averaging the time series of all white matter voxels within a 15mm-radius sphere), 8 RETROICOR time series (4 cardiac, 4 respiration) calculated from the cardiac and respiratory measures taken during the scan (Glover et al., 2000), and 5 Respiration Volume per Time (RVT) time series to minimize end-tidal CO2 effects from deep breaths (Birn et al., 2008). All regressors were detrended with a 4^th^ order polynomial prior to denoising and the same detrending was applied during nuisance regression to the voxel time series. Finally, the residual time series were spatially transformed to standard anatomical space (Talairach-Tournoux).

To ensure that the fMRI data from both groups were high quality and matched, we measured the temporal signal-to-noise-ratio (tSNR) across the whole brain and a summary of in-scanner head motion using the @1dDiffMag program in AFNI. We calculated the tSNR in each voxel as the time series mean divided by time series standard deviation and selected participants from both groups that had high tSNR values across the whole brain. We used Diffmag (comparable to mean Framewise Displacement, Power et al., 2012), which estimates the average of first differences in frame-to-frame motion across each scan run, to exclude participants with scores greater than 0.2 mm/TR. Both tSNR and in-scanner head motion were matched between the groups (Figure 1).

**Figure 1.**
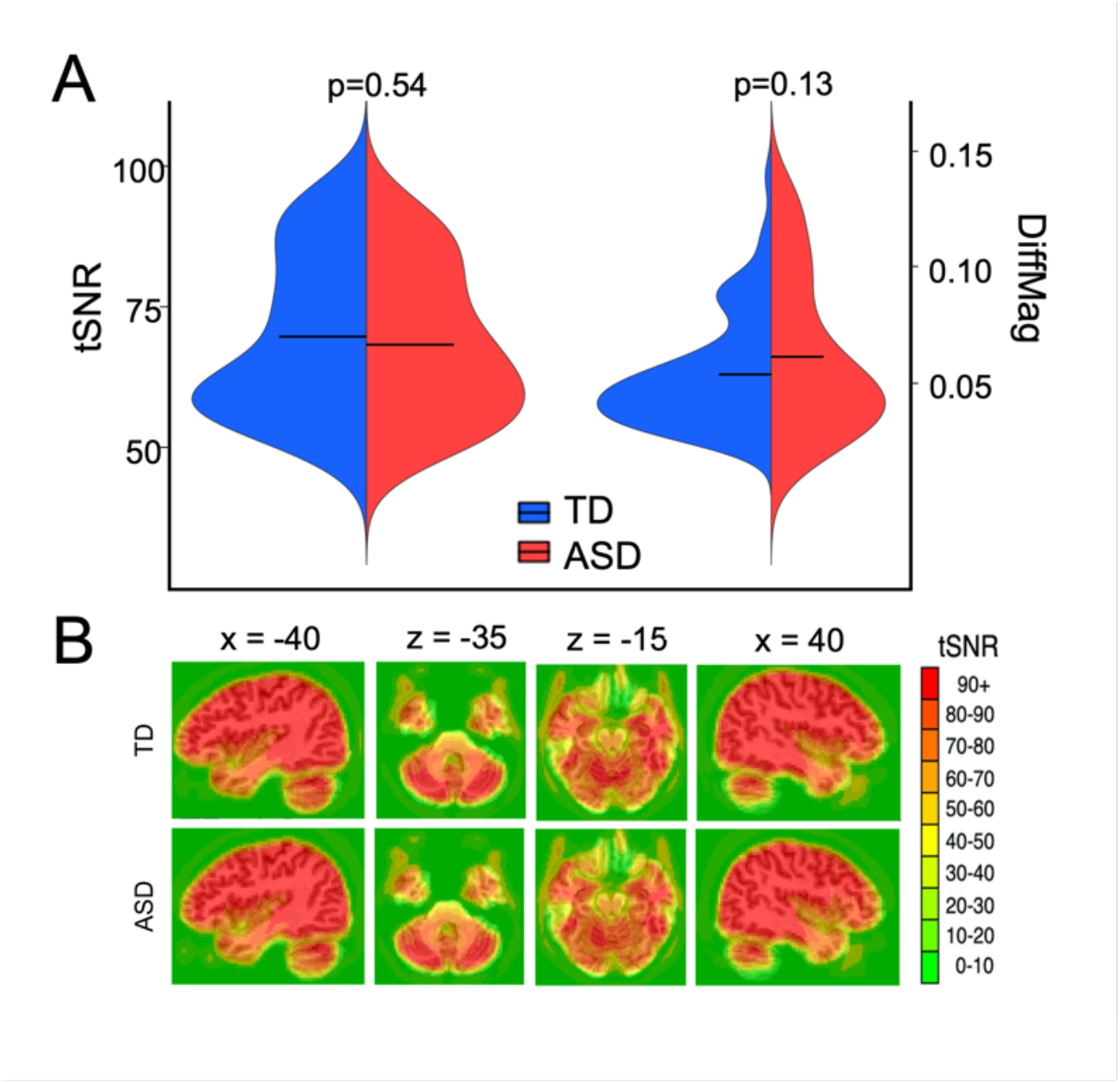
High-quality fMRI data was matched between the groups. **A)** Both groups had high temporal signal-to-noise ratio (tSNR – i.e., time series mean divided by time series SD) and a low level of head motion (as measured using the DiffMag program in AFNI). There were no significant differences between tSNR or DiffMag between the groups. Black horizontal lines in the violin plots represent the mean of each measure in each group. **B)** Whole-brain maps of the average tSNR across participants from the TD and ASD groups, respectively. The tSNR values were high across the whole brain in both groups.

### Alignment of homotopic locations through non-linear registration

Homotopic locations in the two hemispheres were identified and aligned using non-linear volumetric registration with the AFNI 3dQwarp tool (Cox, 1996). Specifically, in each participant, we first flipped the right hemisphere of the anatomical image across the midline so that it was roughly overlapping the left hemisphere. Next, we applied a nonlinear transformation with no blurring to align the right hemisphere with the left. After achieving satisfactory alignment of the right and left hemisphere anatomical images, we applied the same transformation to the resting state data maps. A consequence of this transformation is that right hemisphere voxels were tightly aligned to the corresponding left hemisphere voxels and thus could be projected directly onto the left hemisphere.

### fMRI data analysis: calculating laterality metrics

First, we treated each voxel as a seed, in turn, and correlated (Pearson’s r) the residual EPI time series from the seed with every other voxel in both hemispheres (Figure 2A). The correlations from a seed and all voxels in each hemisphere were averaged separately and stored in the seed voxel. Thus, each voxel was given two values that represented its strength of connectivity within and between hemisphere (Liu et al., 2009; Gotts et al., 2013). For example, each voxel in the left hemisphere was assigned an average correlation with all other voxels in the left hemisphere (Left seed-to-Left hemisphere connectivity was labelled as “LL”) and with all voxels in the right hemisphere (Left seed-to-Right hemisphere connectivity was labelled as “LR”). The same was done for right hemisphere seed voxels (Figure 2A). Next, we applied Fisher’s z-transform to these averaged correlations to yield normally distributed values and then used contrasts to calculate two main forms of functional laterality in each voxel: Segregation and Integration (Figure 2B). Segregation is the tendency for greater within-relative to between-hemisphere interactions and was calculated as (LL– LR)−(RR−RL), thus positive values indicate Left Segregation and negative values indicate Right Segregation. While a large positive value of one side of this equation (e.g., LL−LR) would indicate a stronger average within-than between-hemisphere correlation from a seed voxel, and thus is a measure of laterality, a large value (positive or negative) of the full Segregation metric would indicate that the bias for stronger within-hemisphere interactions is greater for one hemisphere relative to the other. By contrast, the second form of lateralization, Integration, was calculated as (LL+LR)−(RR+RL), thus positive values of the Integration metric indicate relatively stronger bilateral interactions in left-hemisphere voxels and negative values indicate stronger bilateral interactions in the right hemisphere.

**Figure 2.**
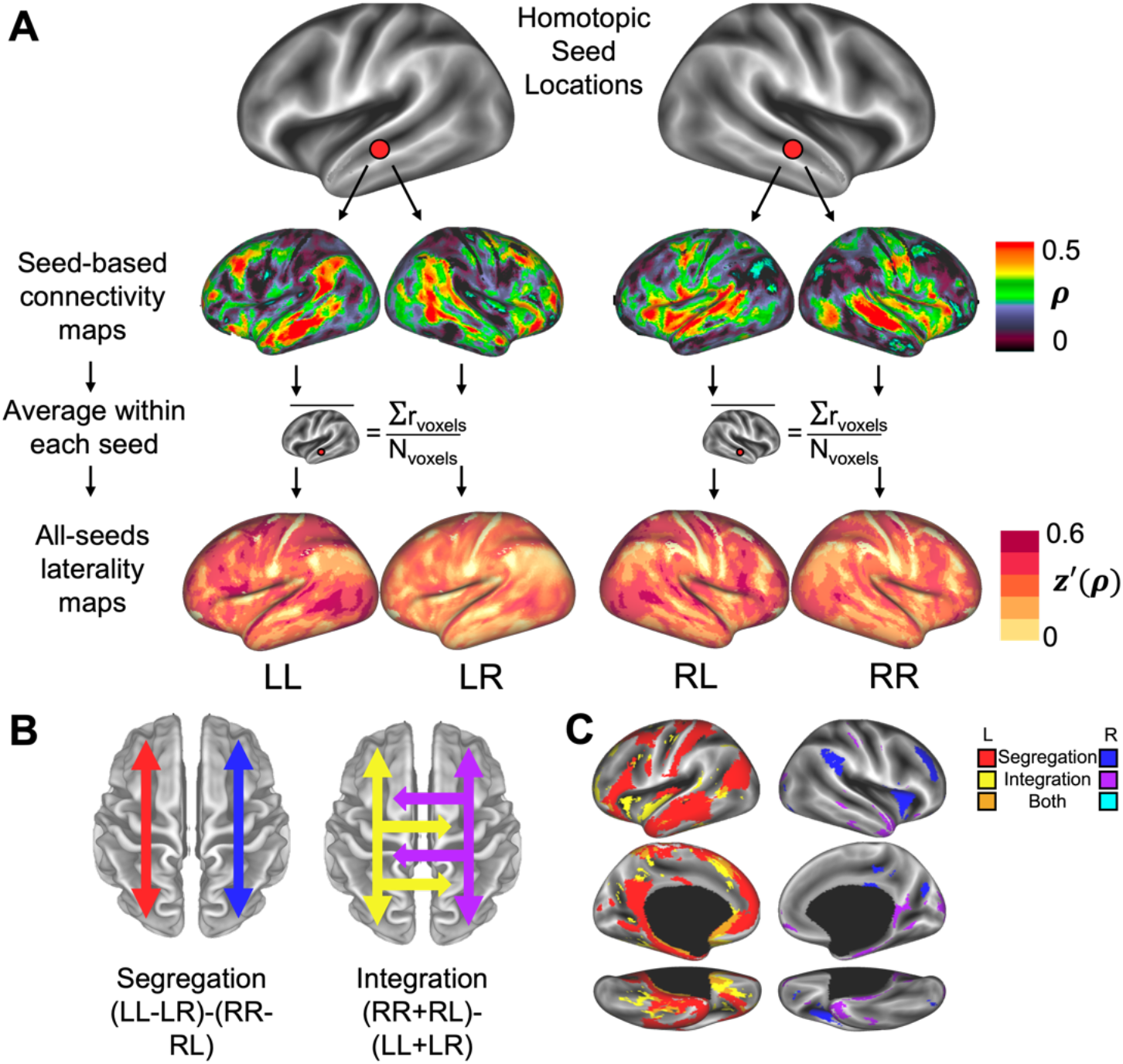
Calculating within- and between-hemisphere connectivity at homotopic locations across the whole brain. **A)** Homotopic voxels in the two hemispheres were identified after aligning the hemispheres using a non-linear transformation procedure and then used as seeds in our laterality calculations (top panel). In every voxel, we correlated the BOLD timeseries with the timeseries from every other voxel to obtain a functional connectivity map in both hemispheres (middle panel – data from an individual participant). We then stored the average correlation coefficient (ρ) from each seed to each hemisphere at the seed voxel location (bottom panel – data from an individual participant), so that all voxels in the brain ended with four values: “LL,” “LR,” “RR,” and “RL.” The first letter in the labels indicates a seed location in the left (L) or right (R) hemisphere, while the second letter indicates the target hemisphere. We then applied the Fisher’s z′-transform to yield normally distributed values and then averaged each brain map across all participants. **B)** After obtaining measures of within- and between-hemisphere connectivity in every voxel in each group, we calculated two main metrics of laterality: one that measures the degree of within-hemisphere communication from each location in cortex relative to the homotopic location in the other hemisphere, referred to as “Segregation,” and another that measures between-hemisphere communication, referred to as “Integration.” **C)** After calculating whole-brain maps of Segregation and Integration in each group, we also made combined-group maps by averaging the data from all of the participants from both groups together and then thresholded the maps (p<0.001). We used the combined-group maps to functionally define networks of interest for our main analyses.

### fMRI data analysis: Identifying functional networks

After calculating the laterality metrics in all voxels, we obtained whole-brain maps of Segregation and Integration for both hemispheres (Figure 2C). We then applied a stringent threshold (p<10^−6^, FDR q<0.01) to the maps, so we could identify individual regions for further analyses. Next, we extracted the average timeseries from each region and correlated it with the timeseries from every voxel in the brain to obtain a whole-brain connectivity pattern for each region. We then correlated the whole-brain patterns of connectivity between the regions and submitted the resultant region x region correlation matrix to k-means clustering. Specifically, the square ROI correlation matrix was iteratively analyzed with k-means cluster analysis at progressively larger numbers of clusters (*k*) and each choice of *k* was repeated 100 times for stability.

## Results

### Functionally defining a network of interest

Before comparing patterns of laterality between ASD and TD individuals, we first defined functional networks by combining the groups (N=140) and identifying brain regions based on four patterns of laterality: Left segregation, Right segregation, Left integration, and Right integration (Figure 2C). We found a total of 18 regions across the brain: eight regions identified with the Left-segregation metric, one with Right-segregation, five with Right-integration, and four with Left-integration (p<10^−6^, FDR q<0.01). We submitted the resultant 18×18 correlation matrix to k-means clustering and found an optimal trade-off of cluster number and variance explained by k-means clustering at the choice of k=3 clusters, with approximately 80% of the variance explained (Figure 3A). Even though the metric used to identify a region did not guarantee that it would be clustered with other regions identified using the same metric, a mostly frontotemporal network comprised seven of the eight regions that were identified using the Left-segregation metric (the red network in Figure 3B) – the inferior frontal gyrus (IFG), (dorsal and ventral) medial prefrontal cortex (mPFC), anterior middle temporal gyrus (MTG)/temporal pole, anterior hippocampus, posterior MTG/superior temporal sulcus (STS), angular gyrus, and posterior cingulate cortex (PCC)/precuneus. This network also included two regions defined with the Right and left Integration metrics, respectively: anterior superior temporal gyrus (aSTG) and lateral anterior temporal pole (Figure 3D). We used this conglomeration of language and social processing regions (Geschwind, 1972; Binder et al., 1997; Frith and Frith, 2007; Olson et al., 2007; Adolphs, 2009; Mitchell, 2009; Fedorenko et al., 2011) as the primary network-of-interest (NOI) for further analyses.

**Figure 3.**
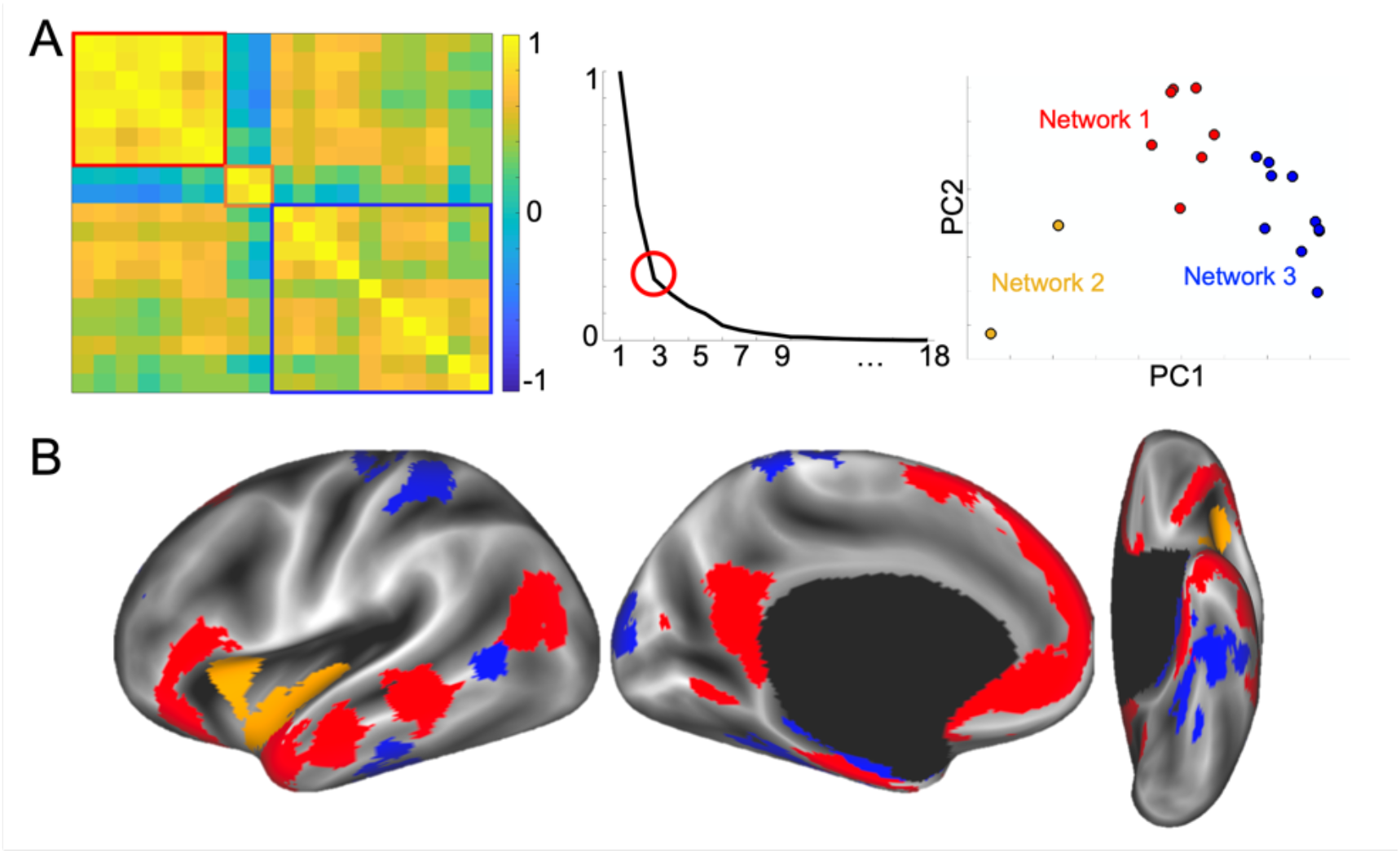
Functionally defined networks of interest. **A)** After we defined 18 regions based on the left and right Segregation and Integration maps, we extracted the timeseries from each, calculated a whole-brain correlation map from each region, then created an 18×18 similarity matrix based on the whole-brain correlation patterns (left panel). Next, we ran k-means clustering on the similarity matrix and found an optimal solution of k = 3 clusters explained roughly 80% of the variance in the data (middle panel). Principal components analysis shows the distribution of the 18 regions separated into three clusters in a 2D plot (right panel). **B)** The three functionally defined networks displayed on an inflated brain. Network 1 (red) is our primary network of interest (NOI), since the regions in it predominantly exhibited strong left Segregation and overlap with previously reported regions related to language and social communication.

### Testing four models of “reduced” laterality in the NOI

We tested four models of “reduced” laterality in the NOI in ASD: A model of increased communication between hemispheres (the Integrated model); A model of no change in the asymmetry of (left) lateralized connectivity between hemispheres (the Left-segregated, or Null, model); A model of decreased laterality in left hemisphere regions accompanied by increased laterality in homotopic regions in the right hemisphere (the Right-segregated model); A model of no change in the typically strong laterality of left hemisphere regions accompanied by increased laterality of homotopic regions in the right hemisphere (the Decoupled-hemispheres model). First, we tested the Integrated model using the Integration metric to ask whether there is more inter-hemispheric communication in the ASD network compared to the TD group. The Integration metric was not significantly different from zero in either group (both t’s_(69)_<1, both p’s>0.4) and there was not a significant difference between the groups (t_(138)_=0.08, p=0.94) (Figure 4A), thus suggesting that “reduced” laterality in the NOI is not due to an increase in communication across the hemispheres. However, it could be the case that in the ASD group there is an increase in the across-hemisphere component metrics (LR and RL), which measure the strength of communication between the typically left-lateralized network and the left and right hemisphere, respectively, separate from the degree of within-hemisphere or homotopic connectivity. If the across-hemisphere metrics are greater in the ASD than TD group, it would suggest that left hemisphere regions in the NOI are communicating more with regions in the right hemisphere. Interestingly, however, we instead found the opposite – i.e., decreases in the LR (t_(138)_=-1.90, p=0.06) and RL (t_(138)_=-2.42, p=0.02) metrics in ASD compared to the TD group (Figure 4C). These results suggest that the typically left-lateralized NOI in ASD is less connected to the right hemisphere, while homotopic right regions are less connected with the left hemisphere.

**Figure 4.**
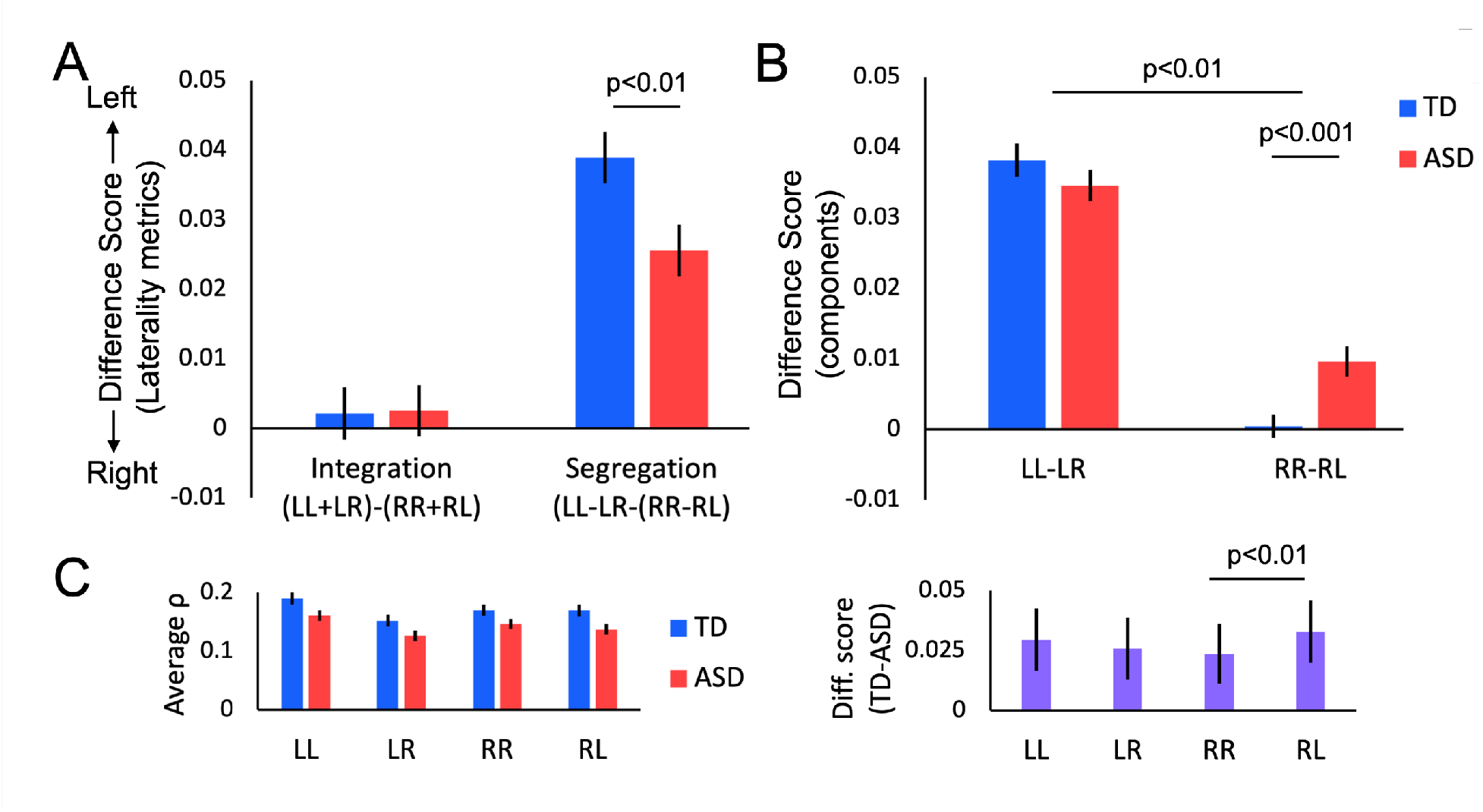
Laterality differences in the NOI. **A)** Integration and Segregation scores averaged within each group. Positive values indicate leftward Integration/Segregation and negative values indicate rightward Integration/Segregation. The Integration metric did not differ from zero in either group nor was there a significant between group difference. By contrast, both groups showed significant left Segregation and there was a significant decrease in left Segregation in the ASD compared to TD group. **B)** Comparison of each side of the Segregation metric separately. Greater values of “LL-LR” indicate stronger laterality of regions in the left hemisphere, while greater values of “RR-RL” indicate stronger laterality of regions in the right hemisphere. There was strong laterality in the left hemisphere of both groups and no significant difference between the groups. In the right hemisphere, the TD group showed no laterality, while there was significant right hemisphere laterality in the ASD group. The difference in right hemisphere laterality was significant between the groups. Furthermore, there was a significant 2 Group x 2 Laterality metric interaction. **C)** The average correlation coefficient for each component metric (e.g., LL = the average correlation between all left hemisphere seeds in the NOI and all left hemisphere voxels) in both groups. The left panel shows the component values separately for each group, while the right panel displays the components as between-group difference scores to make the group differences more apparent. All error bars are +/- 1 standard error from the mean.

Next, we tested the Left-segregated model using the Segregation metric. In the TD group, the NOI exhibited strong leftward segregation (i.e., significantly stronger left laterality compared to the laterality of homotopic right regions), thus we can reject the Left-segregated model if there is a significant decrease in this laterality metric in the ASD group. As predicted, a two-sample t-test revealed a significant decrease in the Left-segregation metric in ASD compared to the TD group (t_(138)_=-2.57, p=0.01), thus suggesting that ASD individuals indeed exhibit atypical left laterality in the NOI (Figure 4A).

Finally, we tested the Right-segregated and Decoupled-hemispheres models by looking at the components of the Segregation metric separately. As described above, the Segregation metric is a measure of the degree of laterality in voxels of one hemisphere relative to the laterality of homotopic voxels in the other hemisphere. Thus, one side of the equation (LL-LR) indicates the degree of left laterality independent of homotopic regions in the right hemisphere, while the other side (RR-RL) indicates the degree of right laterality at the homotopic points, independent of the left hemisphere. If the Right-segregation model is correct, then we should see a significant decrease in left laterality coupled with a significant increase in right laterality in the ASD compared to TD group. By contrast, if the Decoupled-hemispheres model is correct, then we should see no difference in left laterality coupled with a significant increase in right laterality in the ASD compared to TD group. A 2 (Group: ASD, TD) x 2 (Laterality: Left, Right) two-way ANOVA (Figure 4B) revealed a significant interaction (F=9.30, p<0.01) and planned-comparisons t-tests showed that there was not a significant difference in left laterality between the groups (t_(138)_=-1.12, p=0.26), while there was a significant increase in right laterality in the ASD compared to the TD group (t_(138)_=3.40, p<0.001). While these results support the Decoupled-hemispheres model, it could be the case that the difference in right laterality is driven solely by increased connectivity within the right hemisphere (RR) rather than a decrease in connectivity to the left hemisphere (RL) in the ASD group. To ensure that the difference between the groups is due to decreased connectivity between hemispheres in the NOI, we conducted a 2 (Group: ASD, TD) x 2 (Component: RR, RL) two-way ANOVA and found a significant interaction (F=10.19, p<0.01) that was driven by a significantly greater decrease in the RL component in ASD compared to the TD group compared to the decrease in RR (Figure 4C). A two-way ANOVA for the left hemisphere components (LL and LR) did not reveal a significant interaction (F=1.04, p=0.31). Taken together, these results suggest that while the ASD group has an intact left-lateralized network, the homotopic regions in the right hemisphere are more strongly lateralized relative to the TD group.

### Correlations between left and right laterality in the NOI and a measure of verbal ability

Next, we correlated the left- and right-laterality metrics with verbal IQ scores (while covarying the effects of age, block IQ, and head motion) and then compared the correlation coefficients between the groups. We found that verbal ability was positively correlated with the left-laterality metric in both groups (ASD: r=0.38, p=0.002, TD: r=0.26, p=0.03), while verbal ability was negatively correlated with right-laterality in the ASD (r=-0.28, p=0.03), but not TD group (r=-0.002, p=0.99) (Figure 5). Crucially, the negative correlation between verbal ability and the right-laterality metric in the ASD group was significantly different from the same correlation coefficient in the TD group (z=-1.64, p=0.05) and significantly different from the within-group correlation coefficient between verbal ability and the left-laterality metric (z=-2.46, p<0.01). Furthermore, neither left nor right laterality was correlated with block IQ scores, while covarying the effects of age, verbal IQ, and head motion (all r’s<0.08, all p’s>0.50). This pattern of relationships between verbal ability and hemispheric laterality in ASD further supports the Decoupled-hemispheres model and suggests that atypical right laterality of the NOI in ASD is a maladaptive trait.

**Figure 5.**
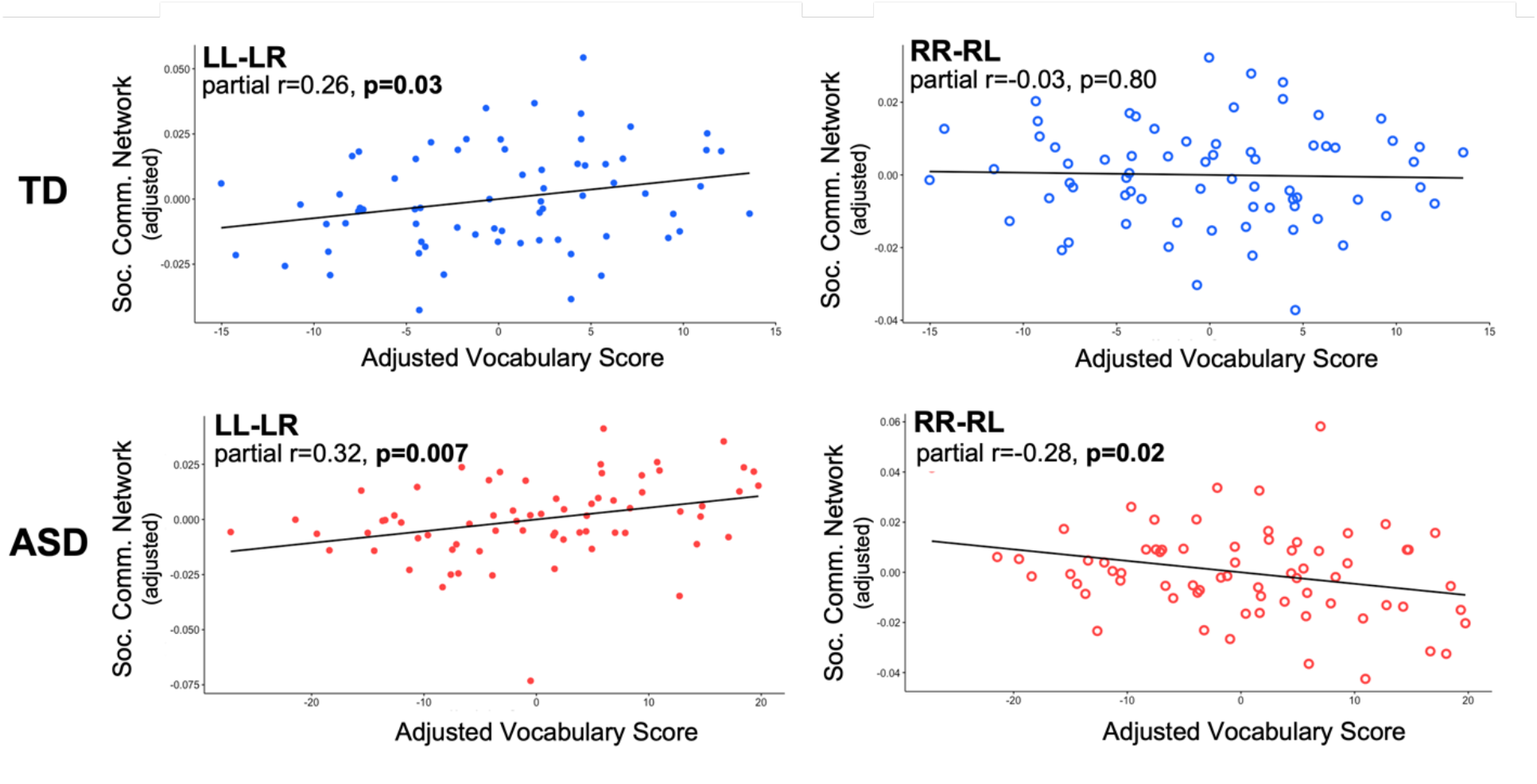
Correlations between laterality measures and verbal ability. In the left panel, both TD and ASD left hemisphere laterality in the NOI is positively correlated with verbal ability. By contrast, right hemisphere laterality and verbal ability are not correlated in the TD group, but there is a negative correlation between the two variables in the ASD group (right panel). The plotted data are correlations of the residuals of the brain and behavior data after regressing out Age, head motion, and Block IQ score.

### Are these patterns of results specific to the NOI in ASD

In addition to the typically left-lateralized NOI that was the focus of our analyses, we also identified two other networks using metrics of laterality and k-means clustering (Figure 3B). One network was composed of regions in the insula and STG that were defined using the Right-segregation and Left-integration metrics. The other network comprised regions in somatosensory, occipital, and dorsal lateral prefrontal cortices, and the parahippocampal gyrus that were defined using the Left and Right integration and Left-segregation metrics. Since a large body of literature has focused specifically on the typically left-lateralized network (comparable to our NOI) in relation to communication-and language-related deficits in ASD, we predicted that the other two networks would not show the same pattern of atypical laterality. As predicted, there were no significant differences between the groups in any of the laterality metrics (all p’s>0.18). Thus, atypical laterality in the ASD group appears to be specific to the (typically left-lateralized) NOI.

## Discussion

We tested four models of “reduced” laterality in a social-communication network in ASD individuals using high-quality fMRI data and a resting-state fMRI method of measuring patterns of functional laterality across the brain. We found that the social-communication network exhibited the typical pattern of left laterality in ASD when compared to a tightly matched TD control group. However, we also found within the same network a significant increase in laterality of homotopic regions in the right hemisphere in ASD individuals. In both groups, left laterality was positively correlated with a measure of verbal ability, while right laterality in ASD, but not TD, was negatively correlated with the same verbal measure. These results suggest that “reduced” laterality of this network in ASD is due to the two hemispheres becoming more independent than seen in typically developing individuals and that the increase in right laterality of this network in ASD is maladaptive. Crucially, these differences in patterns of laterality were not found in two other functional networks and were specifically correlated to a measure of verbal, but not visuospatial, ability.

We designed our experiment to address three shortcomings that are often found in prior studies of differences in patterns of laterality between ASD and TD groups. First, we selected high-quality fMRI data from a relatively large sample of ASD individuals (N=70), as determined by measures of tSNR and head motion (Figure 1), and a tightly matched control group of TD individuals that were also matched on age and full-score IQ. Therefore, we could be confident that differences between our groups were due to functional changes of interest rather than poor data quality or demographic differences. Second, our method allowed us to define functional networks based on patterns of resting-state correlations and then probe the defined networks with independent measures of laterality, thus freeing us from the need to define a priori networks based on anatomy or coordinates from prior studies. Third, we measured changes to the functional organization of a social-communication network in ASD, so we could adjudicate between models of functional laterality that might underlie prior reports of “reduced” laterality in ASD individuals.

While several prior studies have used either task-based fMRI or measures of cortical volume to establish that atypical, or “reduced,” laterality of regions involved in social and communication processes is a stable feature of ASD, it is unclear how the observed changes in laterality are related to the underlying functional organization of the network. In the case of task-based fMRI studies, the evidence for “reduced” laterality is an imbalance in the relative magnitude of responses between the left and right hemisphere regions of the network. For example, several studies have found that during language and communication tasks, the magnitude of responses is significantly greater in the left hemisphere compared to homotopic regions in the right for TD individuals, while in ASD individuals the magnitude of responses is either statistically equal between the hemispheres or even stronger in the right hemisphere (Lindell and Hudry, 2013; Herringshaw et al., 2016). While these results do indeed demonstrate a reduction in the typical left laterality of the network, the functional architecture underlying the reduction is left unknown. Therefore, we used functional connectivity to measure how the organization of the network is changed in ASD. We tested four models of potential changes to the organization of the network that would explain reductions in laterality found using task-based fMRI. Our analyses indicated that while the laterality typically found in left hemisphere regions is intact in ASD individuals, homotopic regions in the right hemisphere are more strongly lateralized in the ASD compared to TD group. These results suggest that “reduced” laterality in the functional networks underlying social and communication processes in ASD individuals is due to the left and right hemispheres decoupling from one another and acting as independent networks. Furthermore, we found that the degree of atypical laterality in right hemisphere regions in ASD individuals is negatively correlated with verbal ability and thus seems to be maladaptive. Intriguingly, these results are consistent with deficits in language and social communication skills related to agenesis of the corpus callosum, a condition that results in reduced communication between hemispheres of the brain (Paul et al., 2003).

In conclusion, we found that previous reports of “reduced” laterality in regions underlying language and social communication in ASD is a result of the hemispheres becoming decoupled and thus behaving more like independent, intrahemispheric networks. We further found that the degree to which right hemisphere regions are lateralized (i.e., decoupled from the left hemisphere) was negatively correlated with verbal ability in ASD. These results offer a detailed account of how patterns of functional laterality shift in a social-communication network in ASD individuals and, we hope, these results may lead to more precise clinical identification and interventions for social-communication deficits in ASD individuals.

## Data Availability

All data produced in the present study are available upon reasonable request to the authors

## Acknowledgments

We thank Greg Wallace for insightful discussions and technical assistance. This work was supported by the NIMH Intramural Research Program. (#ZIA MH002920-09, clinical trials number NCT01031407). The authors declare no competing financial interests.

